# The Role of Neutrophil PD-L1 Expression in Acute Exacerbation of COPD

**DOI:** 10.1101/2025.11.23.25340811

**Authors:** Wei Sun, Liyu Zheng, Haibin Li, Li An, Jing Wang, Chao Ren

## Abstract

**Background and Objective:** The elevation of neutrophil-lymphocyte ratio (NLR) has been shown to be critically involved in unfavorable outcomes among patients with chronic obstructive pulmonary disease (COPD). While the mechanism underlying the changes of NLR during COPD progression remains unclarified.

**Methods:** In this prospective observational study from June to December 2024, we included 14 patients with acute exacerbation, 27 with stable conditions, and 9 controls. Blood samples were collected for neutrophil isolation, and the CD15^+^CD274^+^ neutrophil ratio was measured using flow cytometry. We also recorded and analyzed smoking history, pulmonary function test results, and blood routine test data.

**Results:** The ratio of CD15^+^CD274^+^ neutrophils was notably higher in patients with COPD compared to healthy controls and was even more elevated in those experiencing acute exacerbations of COPD (AECOPD) than in patients with stable COPD. This increased ratio was positively correlated with NLR but negatively correlated with FEV1% (FEV1 percentage of the predicted value). The CD15^+^CD274^+^ neutrophil percentage is a powerful biomarker for predicting AECOPD (AUC 0.894; 95% CI 0.8–0.989). A ratio above 3.273 (odds ratio 1.386; 95% CI 1.016–1.891; P=0.039) independently predicts AECOPD.

**Conclusion:** The increased ratio of CD15^+^CD274^+^ neutrophil serves as an independent risk factor in predicting the incidence of AECOPD, which might be due to disrupting NLR and worsening FEV1%. Elevated PD-L1(programmed death ligand 1, CD274^+^) on neutrophils might also be a potential therapeutic target for reversing immune dysfunction and poor outcomes of patients with COPD.

**Key messages:** **What is already known on this topic** – An increased neutrophil-lymphocyte ratio (NLR) is linked to poor outcomes in COPD patients, though the reasons for NLR changes during COPD progression are still unclear.

**What this study adds** – Higher level of CD15^+^CD274^+^ (PD-L1) neutrophil, showing a positive correlation with NLR and a negative correlation with FEV1%, serves as a strong biomarker for predicting AECOPD.

**How this study might affect research, practice or policy** – Elevated CD15^+^CD274^+^ neutrophil may indicate COPD progression by suppressing lymphocyte activity, causing abnormal NLR changes. Targeting elevated PD-L1 on neutrophils may improve immune function and outcomes in COPD patients.

## Introduction

Chronic obstructive pulmonary disease (COPD) is associated with chronic inflammatory response which is marked by an elevated presence of various immune cells and disturbed responses(1). The loss of balance between innate and adaptive immune response, such as the ratio of neutrophil-to-lymphocyte (NLR), is one of the major signs of immune dysfunction and critically involved in the development of various diseases(2, 3). Notably, patients with COPD show marked activation of neutrophils and suppression of lymphocyte function(4). The NLR has been well investigated in numerous studies as a predictive marker for disease severity(5), hospitalization(6), and mortality(7) among COPD patients. However, the underlying mechanisms remain incompletely understood.

Previous research has identified that the interaction between programmed death receptor 1 (PD-1) and its ligand (PD-L1) is an important inhibitory immune checkpoint, which has been reported by microorganisms and tumors to attenuate antimicrobial or tumor immunity and facilitate chronic infection and tumor survival. CD15 marks neutrophils, and CD274 indicates PD-L1 expression; thus, CD15^+^CD274^+^ neutrophils signify PD-L1-expressing neutrophils. The expression of PD-L1 can be modulated by a variety of factors, including pharmacological agents(8), persistent infections(9), and hypoxic conditions(10). Among patients with sepsis, there is an upregulation of PD-L1 expression on neutrophils, which induces lymphocyte apoptosis through direct interaction(11). Among patients with COPD, studies have demonstrated a decrease in PD-L1 expression levels on the surface of macrophages and dendritic cells(12, 13), hinting the critical involvement of PD-L1 in regulating prognosis of COPD patients by regulating host immune response. PD-L1 is a key immune checkpoint, and its abnormal levels indicate immune dysfunction. Assessing immune function in COPD patients is vital and linked to future targeted drug treatments. There are no studies to date that have investigated whether PD-L1 expression on neutrophils directly correlated with NLR alterations or exacerbations risk in patients with COPD. Our prior research(14) indicates a significant association between the NLR and poor prognosis in patients with AECOPD (acute exacerbation of COPD). A comprehensive review of the literature reveals that neutrophils can suppress lymphocyte count and function via their surface expression of programmed death-ligand 1 (PD-L1)(11), thereby influencing NLR levels. This study aims to evaluate the potential role of neutrophil PD-L1 as a risk factor for AECOPD and to investigate the correlation between neutrophil PD-L1 expression and the NLR (Figure 1).

**Figure 1.**
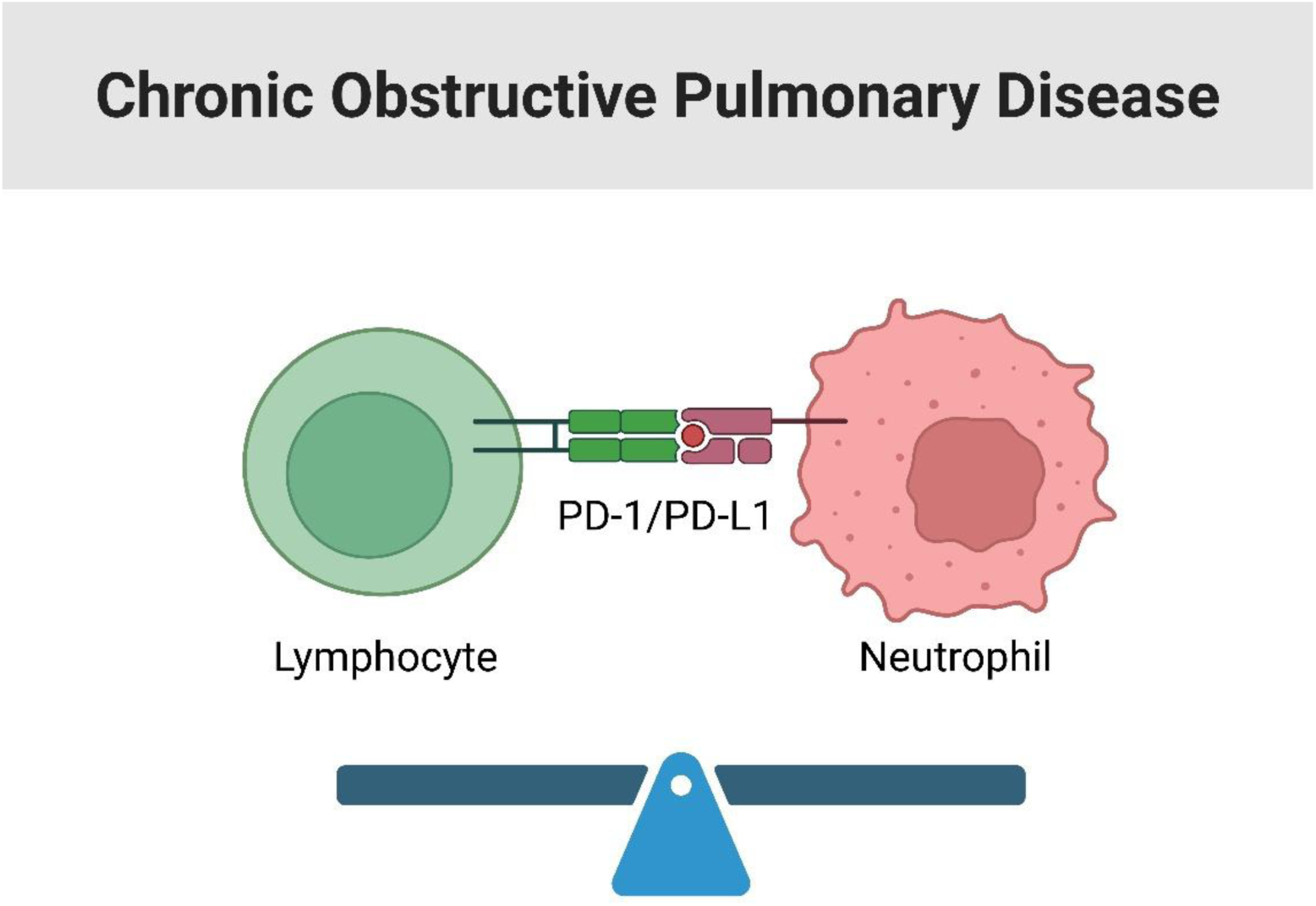
Among patients with COPD, the expression of PD-L1 shows significant upregulation on the surface of neutrophil, which may inhibit lymphocyte proliferation and function through interaction with PD-1 on lymphocyte. This interaction might be major cause for disturbed balance of NLR, resulting in the acute exacerbation of COPD. **Abbreviations:** COPD=chronic obstructive pulmonary disease; NLR=neutrophil/lymphocyte ratio, PD-1-PD-L1=programmed death receptor 1 and its ligand.

## Materials and Methods

This study was approved by the ethics committee of Beijing Chao-Yang Hospital(2024-KE-414-1) and informed consent was obtained from all participants. This analysis comprises data from a total of 41 patients with confirmed COPD, including stable COPD patients (n=27) and those with acute exacerbation (n=14). In addition, a total of 9 health control was also included between Jun 2024 and Dec 2024. The subjects participating in this study self-reported as Asian Han Chinese. Patients or the public were not involved in the design, or conduct, or reporting, or dissemination plans of our research.

The detailed inclusion criteria are listed as follow: age≥40 years old; COPD is defined as heterogeneous lung condition characterized by chronic respiratory symptoms (dyspnea, cough, expectoration and/or exacerbations) due to abnormalities of the airways (bronchitis, bronchiolitis) and/or alveoli (emphysema) that cause persistent, often progressive, airflow obstruction(post-bronchodilator FEV_1_/FVC[Forced Expiratory Volume in the 1st second/Forced Vital Capacity] ratio less than 70%) (15). AECOPD is defined as an event characterized by increased dyspnea and/or cough and sputum that worsen in <14 days which may be accompanied by tachypnea and/or tachycardia(16) (Patients with COPD in the acute phase did not undergo pulmonary function tests; therefore, we incorporated the results from prior pulmonary function assessments.). The study included patients with moderate acute exacerbations treated as outpatients, receiving short-acting bronchodilators, oral corticosteroids, and antibiotics. The patients with COPD in this study received inhaler triple therapy. Stable patients experienced no exacerbation for a minimum duration of three months.

Exclusion Criteria: incomplete clinical data; presence of other severe pulmonary diseases (e.g., asthma, bronchiectasis); end-stage of chronic disease (e.g., chronic kidney failure, chronic heart failure and malignancy), patients received immune-associated therapies, including immune checkpoint inhibitors, long-term (>14 days) use of glucocorticoids, and immunosuppressors, patients during pregnancy or lactation.

Patients’ baseline characteristics were recorded, including gender, age, smoking history and comorbidities. The control group was matched with COPD patients based on age, gender, comorbidities, and smoking status. The values of FEV1% (FEV1 percentage of the predicted value), FEV1/FVC and the presence of respiratory failure were recorded among patients with COPD. The values of C-reactive protein (CRP) and routine blood parameters were obtained. NLR was calculated by the ratio of the total number of neutrophils to the total number of lymphocytes.

### Blood neutrophil isolation

A total of 5 ml blood samples were drawn from elbow veins, and further collected in anticoagulant tubes (TBD sciences, China). The blood samples were added into the isolation buffer with volume ratio of 1:1, and then gradients were separated by centrifugation at 600g for 30 min at room temperature with no break. Subsequently, two layers of cells, including the mononuclear cells in the upper layer and granulocytes in the lower layer, were separated. The granulocytes were carefully transferred to a new tube and washed twice by phosphate buffer (PBS) using centrifugation. These neutrophils were counted and resuspended in PBS (3×10^5^/100μL) for flowcytometry staining.

The neutrophils received antibody blocking by 1% bull serum albumin (BSA, in PBS) for 1h at room temperature. Then these cells were stained with CD15 and CD274 by adding FITC-conjugated anti-human CD15 antibody as well as PE-conjugated anti-human CD274 antibody for 1h at room temperature. The ratio of CD15^+^CD274^+^ neutrophils were measured by flow-cytometry analysis (BD Biosciences, USA).

This study aims to investigate the impact of PD-L1 expression on neutrophils in relation to the risk of AECOPD. Currently, there is insufficient information in the literature to accurately determine the necessary sample size for this investigation. Therefore, it is essential to conduct a pilot study to explore the role of PD-L1 on neutrophils in predicting AECOPD.

## Statistical Analysis

To compare categorical variables, the Chi-square test was used. Student’s t-test or Mann–Whitney U-test was used for comparing continuous variables. Pearson correlation was employed to evaluate the relationship between CD15^+^CD274^+^ neutrophil and FEV_1_, and the results were displayed as correlation coefficients and P values. The partial correlation method assessed the relationship between CD15^+^CD274^+^ neutrophil and NLR, controlling for neutrophil count, and results were displayed as correlation coefficients and P values. Receiver-operating characteristic (ROC) curves was established to evaluate the ability of markers for predicting AECOPD. For each ROC curve, the optimal cutoff values, sensitivity, specificity, positive/negative predictive value, diagnostic accuracy, Youden ’ s index, area under curve (AUC), and 95% CI(confidence interval) were calculated. To determine if any markers were independently linked to acute exacerbation of COPD, we used logistic regression with a conditional forward stepwise model, yielding adjusted odds ratios (OR, 95% CIs). Analyses were two-tailed, with significance set at P<0.05.

## Results

There were no significant differences in terms of age, sex, hypertension, coronary heart disease, diabetes and smoking status between patients with COPD and control population. Besides, there were no significant differences in terms of CPR, blood routine and NLR between two groups. Importantly, the percentage of CD15^+^CD274^+^ neutrophil (**Figure 2**) was much higher in patients with COPD than that in control group (**Table 1**).

**Figure 2.**
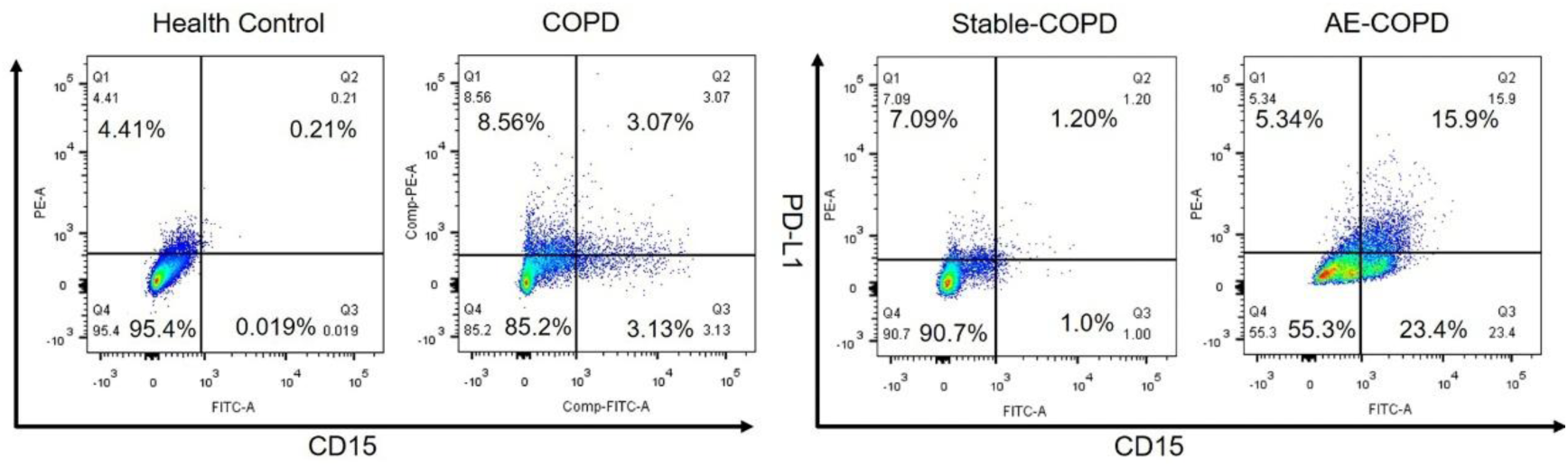
The percent of CD15^+^CD274^+^ cells by flow cytometry **Abbreviations:** Q1: CD15^-^/PD-L1^+^; Q2: CD15^+^/PD-L1^+^; Q3: CD15^-^PD-L1^-^; Q4: CD15^+^PD-L1^-^.

**Table 1.** Comparison between COPD population and health control.

| Characteristics | COPD (N=41) | control<br>subjects(N=9) | P |
| --- | --- | --- | --- |
| Age(years) | 67±10 | 66±10 | 0.849 |
| Sex (male,%) | 36(87.8%) | 7(77.8%) | 0.595 |
| Hypertension(n,%) | 27(65.9%) | 7(77.8%) | 0.699 |
| Coronary heart disease(n,%) | 26(63.4%) | 7(77.8%) | 0.699 |
| Diabetes(n,%) | 31(75.6%) | 6(66.7%) | 0.679 |
| Smoker(current/ex) | 32(41%) | 7(77.8%) | 1 |
| CRP (mg/L) | 4.8(1.2~11) | 5(2.65~5.75) | 0.93 |
| Leukocyte count($\times 10^9/L$) | 6.9±1.65 | 6±1.46 | 0.136 |
| Hemoglobin(g/L) | 139.39±19.59 | 132.89±12.36 | 0.347 |
| Platelet count($\times 10^9/L$) | 209.34±57.05 | 189.33±39.91 | 0.324 |
| Neutrophil count($\times 10^9/L$) | 4.56±1.53 | 3.79±1.57 | 0.178 |
| Lymphocytes count($\times 10^9/L$) | 1.66±0.63 | 1.55±0.32 | 0.621 |
| NLR | 2.57(1.79~4.29) | 2.48(1.39~3.19) | 0.426 |
| Eosinophils count($\times 10^9/L$) | 0.14(0.07~0.24) | 0.15(0.06~0.25) | 0.8 |
| CD15<sup>+</sup>CD274<sup>+</sup> Neutrophil (%) | 3.67(1.08~9.87) | 0.01(0~0.20) | <0.001 |

**Abbreviations:** COPD= chronic obstructive pulmonary disease; CRP= C-reactive protein;
NLR= neutrophil/lymphocyte ratio.

We conducted a comparative analysis of the demographic characteristics and laboratory indices between patients experiencing AECOPD and those in a stable condition (**Table 2**). It showed that the age of AECOPD group was higher than that of stable-COPD. There were no differences in terms of sex, hypertension, coronary heart disease, diabetes and smoking history between two groups. The incidence of respiratory failure was significantly higher in AE-COPD compared with the stable-COPD. FEV_1_/FVC and FEV_1_% were much lower in AECOPD compared with those in stable-COPD. It was also showed that CRP and NLR were higher in AECOPD group than those in stable-COPD, and the levels of hemoglobin, platelet and lymphocytes counts were significantly lower in patients with AECOPD. There were no differences in terms of leukocyte and eosinophils count between AECOPD and stable-COPD patients. The percentage of CD15^+^CD274^+^ neutrophil was significantly higher in AECOPD patients than that in patients with stable-COPD.

**Table 2.**
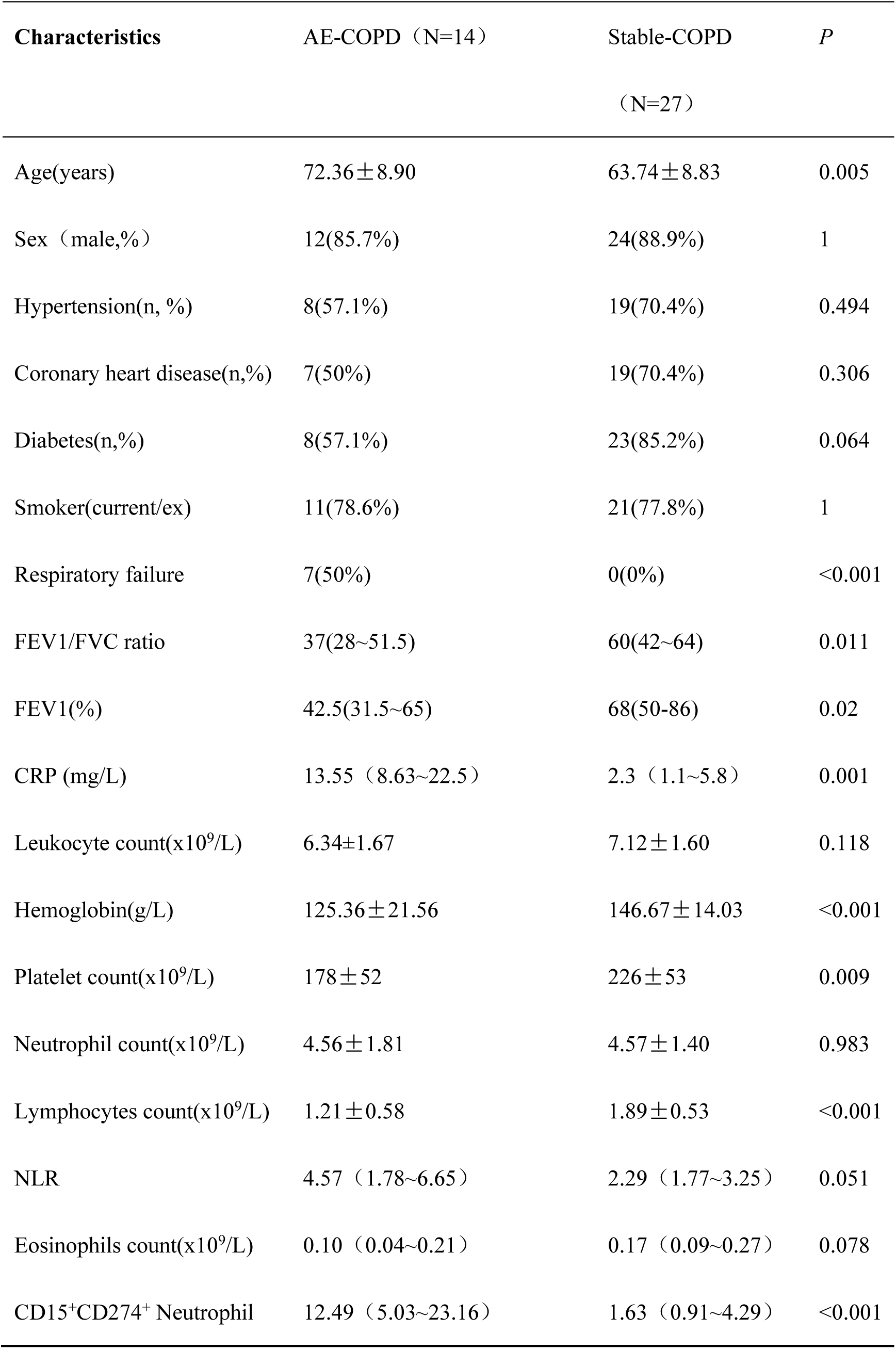
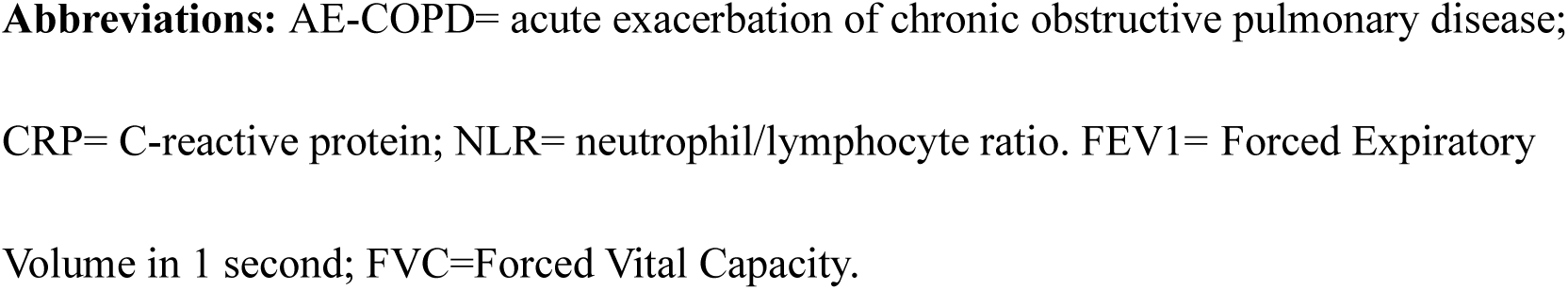
Comparison between AE-COPD patients and stable-COPD patients.

| Characteristics | AE-COPD (N=14) | Stable-COPD<br>(N=27) | P |
| --- | --- | --- | --- |
| Age(years) | 72.36±8.90 | 63.74±8.83 | 0.005 |
| Sex (male,%) | 12(85.7%) | 24(88.9%) | 1 |
| Hypertension(n, %) | 8(57.1%) | 19(70.4%) | 0.494 |
| Coronary heart disease(n,%) | 7(50%) | 19(70.4%) | 0.306 |
| Diabetes(n,%) | 8(57.1%) | 23(85.2%) | 0.064 |
| Smoker(current/ex) | 11(78.6%) | 21(77.8%) | 1 |
| Respiratory failure | 7(50%) | 0(0%) | <0.001 |
| FEV1/FVC ratio | 37(28~51.5) | 60(42~64) | 0.011 |
| FEV1(%) | 42.5(31.5~65) | 68(50-86) | 0.02 |
| CRP (mg/L) | 13.55 (8.63~22.5) | 2.3 (1.1~5.8) | 0.001 |
| Leukocyte count(x10<sup>9</sup>/L) | 6.34±1.67 | 7.12±1.60 | 0.118 |
| Hemoglobin(g/L) | 125.36±21.56 | 146.67±14.03 | <0.001 |
| Platelet count(x10<sup>9</sup>/L) | 178±52 | 226±53 | 0.009 |
| Neutrophil count(x10<sup>9</sup>/L) | 4.56±1.81 | 4.57±1.40 | 0.983 |
| Lymphocytes count(x10<sup>9</sup>/L) | 1.21±0.58 | 1.89±0.53 | <0.001 |
| NLR | 4.57 (1.78~6.65) | 2.29 (1.77~3.25) | 0.051 |
| Eosinophils count(x10<sup>9</sup>/L) | 0.10 (0.04~0.21) | 0.17 (0.09~0.27) | 0.078 |
| CD15<sup>+</sup>CD274<sup>+</sup> Neutrophil | 12.49 (5.03~23.16) | 1.63 (0.91~4.29) | <0.001 |

**Abbreviations:** AE-COPD= acute exacerbation of chronic obstructive pulmonary disease; CRP= C-reactive protein; NLR= neutrophil/lymphocyte ratio. FEV1= Forced Expiratory Volume in 1 second; FVC=Forced Vital Capacity.

Among patients with AECOPD and stable-COPD, the level of NLR was higher and the FEV1% was lower as the percentage of CD15^+^CD274^+^ neutrophil increased (**Figure 3**).

**Figure 3.**
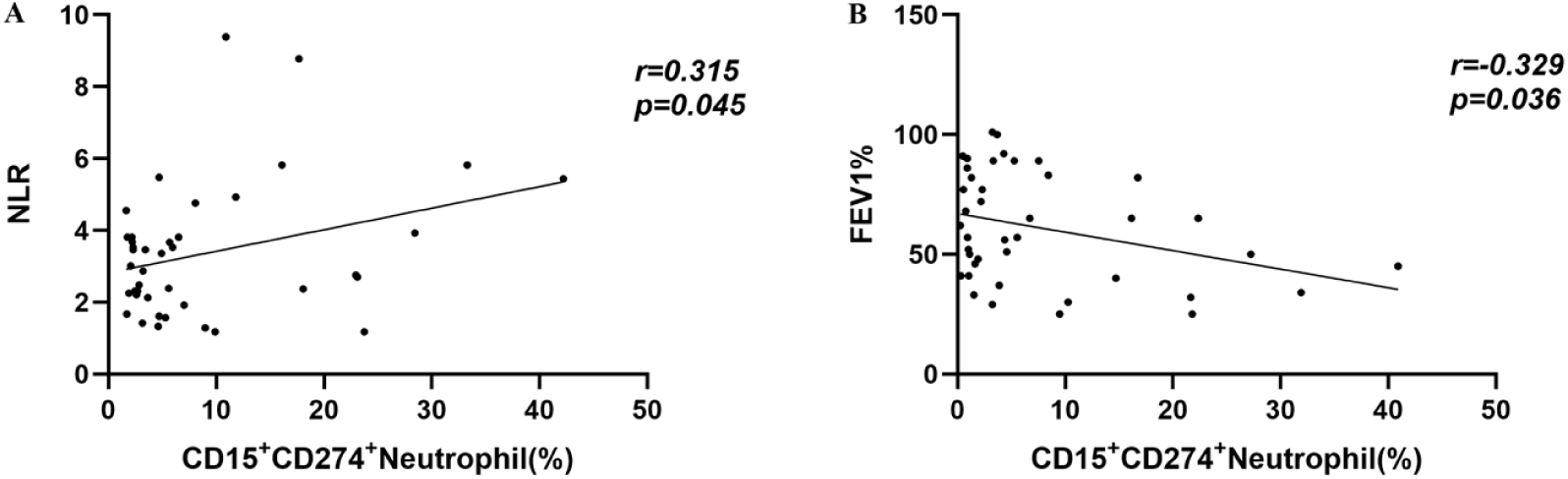
Correlations between the percentage of CD15^+^CD274^+^ neutrophil and NLR (A), and FEV1% (B). **Abbreviations:** NLR= neutrophil/lymphocyte ratio.

The ROC curves of the age, CRP, NLR, CD15^+^CD274^+^ neutrophil, hemoglobin and platelet were displayed in **Figure 4**. **Table 3** showed that the AUC of the percentage of CD15^+^CD274^+^ neutrophil (0.894; 95% CI, 0.8–0.989) was higher than that of age (0.739; 95% CI, 0.568–0.911), CRP (0.815; 95% CI, 0.642-0.988), NLR (0.688; 95% CI, 0.478-0.897), HGB (0.798; 95% CI, 0.638-0.958) and PLT (0.713; 95% CI, 0.55-0.876).

**Figure 4.**
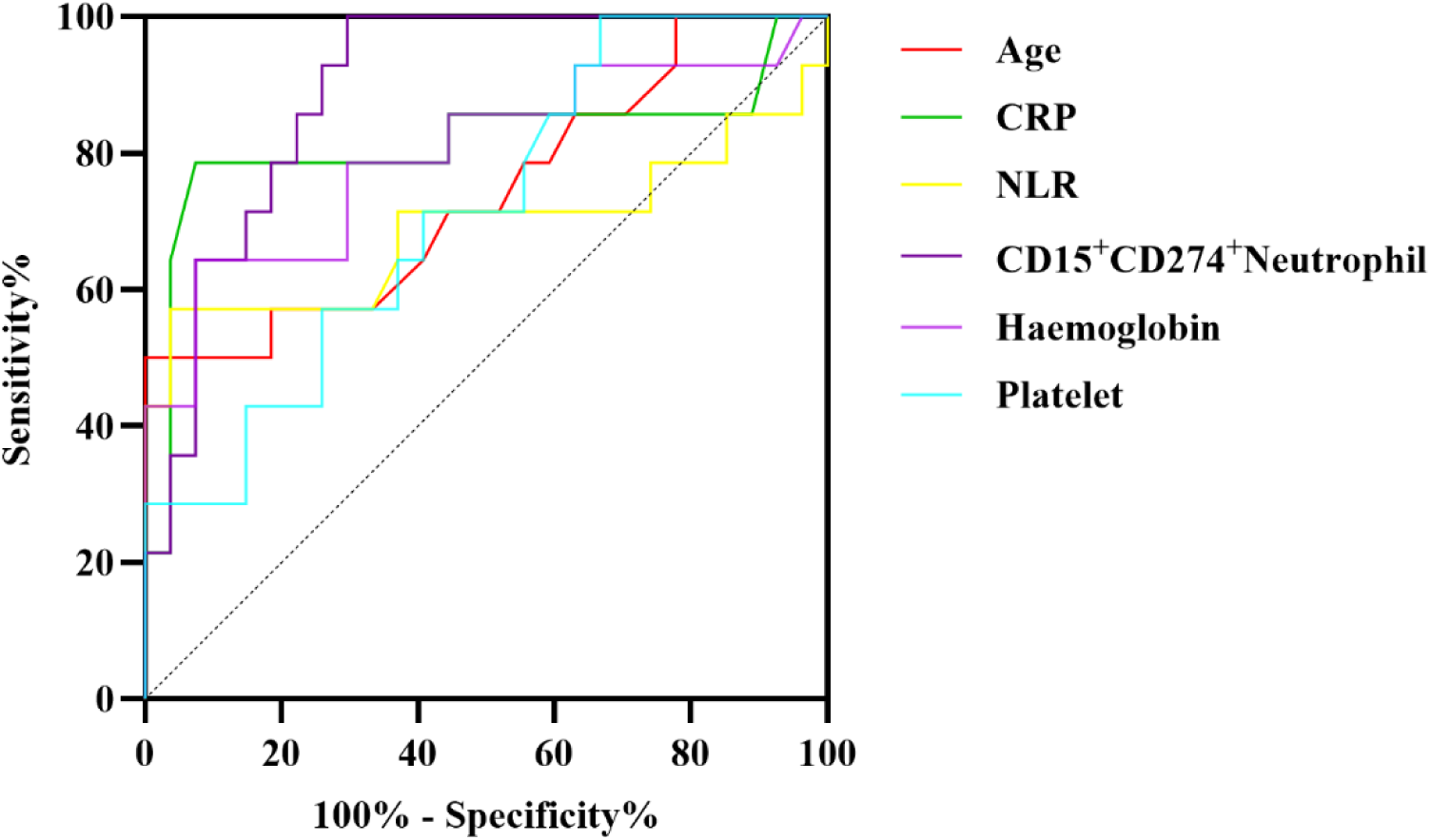
Receiver-operating characteristic curves for age, CRP, NLR, CD15^+^CD274^+^ neutrophil, hemoglobin and platelet for predicting acute exacerbation of COPD. **Abbreviations:** CRP= C-reactive protein; NLR= neutrophil/lymphocyte ratio.

**Table 3.**
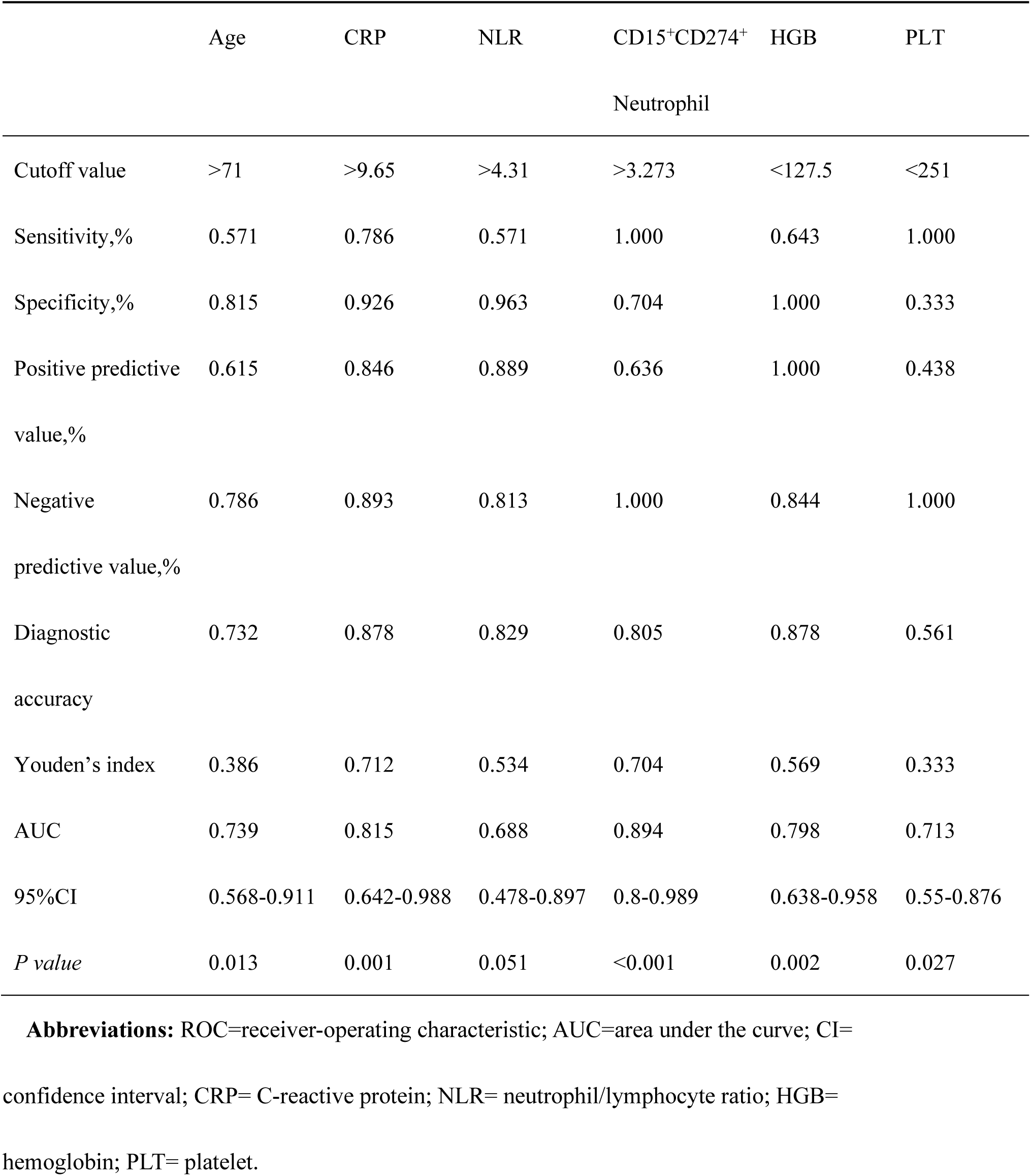
ROC Curve Data.

The diagnostic accuracy of age, CRP, NLR, CD15^+^CD274^+^ neutrophil, hemoglobin and platelet were 0.732, 0.878, 0.829, 0.805, 0.878 and 0.561. The cutoff value for predicting acute exacerbation of COPD was age>71, CRP>9.65, NLR>4.31, CD15^+^CD274^+^ neutrophil>3.273, HGB<127.5 and PLT<251.

In the multivariate logistic regression analysis, we incorporated those factors that exhibited significant differences between the AE and stable groups, while excluding those that did not demonstrate such variations. According to multivariate logistic regression analyses, the percentage of CD15^+^CD274^+^ neutrophil (OR, 1.386; 95% CI, 1.016–1.891; P=0.039) was the only independent factor for predicting acute exacerbation of COPD (**Figure 5**). Multivariate logistic regression analyses were made with covariates including PLT, HGB, NLR, CRP and age.

**Figure 5.**
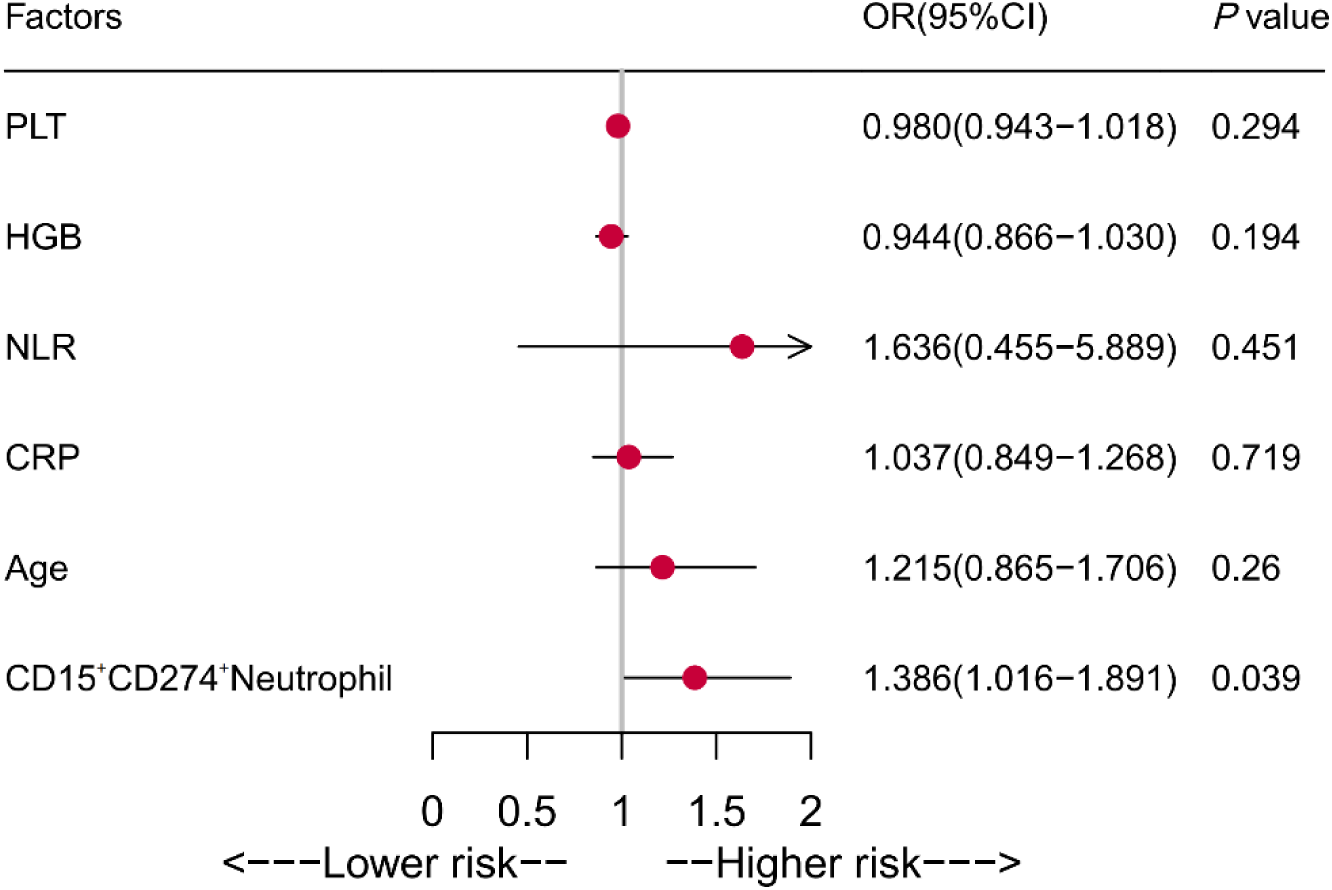
Logistic Regression Analysis of Risk Factors for acute exacerbation of COPD **Abbreviations:** OR, odds ratio; CI, confidence interval; CRP= C-reactive protein; NLR= neutrophil/lymphocyte ratio; HGB= hemoglobin; PLT= platelet.

## Discussion

Our study demonstrated a significant upregulation of PD-L1 expression on peripheral blood neutrophils in patients with COPD, which was correlated with the dysregulation of the neutrophil-to-lymphocyte ratio. Furthermore, elevated PD-L1 expression on neutrophil emerged as an independent risk factor for predicting acute exacerbations of COPD. These findings suggest that increased expression of PD-L1 of neutrophils may play a crucial role in poor outcomes among patients with COPD by mediating NLR imbalance, hinting the importance of immunomodulation on the prognosis of COPD.

The innate immune system meticulously modulates the magnitude, nature, and duration of adaptive immune responses through a variety of mechanisms, including antigen presentation, cytokine secretion, co-stimulatory signaling, and regulatory cell activity (17). The immunopathology of COPD is intrinsically linked to both innate and adaptive inflammatory immune responses. These responses are primarily triggered by the chronic inhalation of cigarette smoke and byproducts from biomass fuel combustion, as well as by intermittent acute exacerbations resulting from infections. This exposure leads to the activation of dendric cells, macrophages, neutrophils and airway epithelial cells. Upon activation, these innate immune cells further facilitate the recruitment and activation of adaptive immune cells by secreting inflammatory mediators(1).

There is no doubt that the imbalance between innate and adaptive immune responses is of great significance yet with predictive values among patients with COPD. The number of macrophages in the small airways increases as the severity of COPD progresses when compared with those of health control (18, 19) along with significantly altered macrophage polarization(20). In addition, there showed retarded maturation of DCs (dendric cell) in COPD, which was also related to disease severity(21). It has been reported that neutrophils presented with excessively activation during COPD by releasing elastase(22), proteinase-3(23), matrix metalloproteinase(24), and myeloperoxidase(25), which was directly involved in the pathophysiology of COPD.

The dysregulation of adaptive immunity is also noteworthy and critically involved in COPD prognosis, which showed weak immune response to pathogens due to dysfunction of effector cells and polarization of suppressive T-regulatory cells (26). The adaptive immune system may also be linked to increased exacerbation risk in the ’frequent exacerbator’ COPD sub-phenotype, characterized by reduced number of CD4^+^ T central memory and CD8^+^ T effector memory cells(27). Yusuke Abe demonstrated that lymphocytes from patients with COPD failed to recognize the nontypeable Haemophilus influenzae(28). In addition, the decreased number of lymphocytes served as an independent risk factors for mortality among COPD patients(29) and was linked to reduced FEV1 levels, poor performance in six-minute walk distances, and worse quality life(30). Therefore, clarifying the crosstalk and its mechanism between innate and adaptive immune responses are of great importance in understanding the progression of COPD as well as providing potential therapeutic targets.

Neutrophil/lymphocyte ratio (NLR)is a potential laboratory indicator with significant implications in predicting all-cause mortality among patients with COPD (31). Our research group has previously demonstrated that NLR was more sensitive to predict noninvasive mechanical ventilation failure than traditional inflammatory markers in AECOPD patients(14). The observed imbalance between the overactivation of neutrophils and the inhibition of T lymphocytes also highlighted the potential link between neutrophils and T cells in COPD patients. The inhibition of lymphocytes is reportedly due to several factors: the increased cortisol levels by intense inflammatory impaired lymphocyte function(32); reduced lymphocyte count due to elevated interleukin-6 (IL-6) levels (33);as well as commonly noted malnutrition status among COPD patients(34). In this study, the increased expression of PD-L1 on neutrophils was noted as an independent risk factor for AECOPD, which might be achieved by significantly elevated NLR.

PD-L1-PD-1 axis delivers inhibitory signals which is responsible for retarded T cell activation and proliferation. The expression of PD-L1 has been identified on murine T and B lymphocytes, dendritic cells, macrophages, mesenchymal stem cells, and mast cells derived from bone marrow(35). Upon ligand binding, PD-1 undergoes phosphorylation at its two intracellular tyrosine residues, and ultimately negatively affect cellular proliferation and cytokine production(36). PD- 1-PD-1 axis plays crucial role in modulating immune defenses against pathogens in response to both acute and chronic infections. It has been reported that the antagonism of PD-1 induced proliferation of effector T cells during adenovirus infection and resulted in rapid viral clearance(37). In mice model with persistent viral infection and CD4^+^T cell dysfunction, blocking the PD-L1/PD-1 pathway benefited CD8^+^T cells by restoring proliferation, cytokine secretion, cell-killing ability, and then reducing viral load(38). In addition, the PD-L1-PD-1 pathway may also be crucial in sustaining chronic bacterial infections. Under exposure to Helicobacter pylori, the expression of PD-L1 significantly increase on human gastric epithelial cells, while anti-PD-L1 antibodies boost T cell proliferation and interleukin-2 (IL-2) production(39).

COPD is characterized by chronic inflammation, which is amplified and further increased during acute exacerbations mostly triggered by infection. Previous study confirmed that patients with COPD showed significantly exaggerated exhaustion of effector T cells due to increased PD-1 expression when compared with healthy controls(40). Anti-PD-1 rescued lung damage and neutrophilic inflammation in COPD mice models(41). The current study showed that the levels of PD-L1 on neutrophil was much higher in patients with COPD than that of control group with no significant differences in terms of the comorbidity and cigarette exposure.

However, our study found no increase in the NLR of patients with COPD compared to healthy individuals, which may be due to the study was limited by a small sample size of patients with COPD, and also with more patients in a stable condition. Study by Ersin showed that NLR was significantly higher in patients with COPD than controls, but not between patients with stable COPD and with exacerbations(42). These differences across studies might result from the included sample sizes and the distribution of patients among different groups. A previous review indicated that lung cancer patients with COPD benefit from anti-PD-1/PD-L1 therapy, with improved lung function, such as FeNO (exhaled nitric oxide) levels, FEV_1_, and FVC, partly due to increased PD-1 expression on CD8^+^ cells among patients with COPD (43). Currently, there is no available immunotherapy for COPD, therefore, further exploration and research in this area may be warranted in the future.

In current study, compared with stable COPD, patients with AECOPD exhibited higher NLR and we further confirmed that neutrophil PD-L1 expression was positively correlated with NLR. However, no significant differences in NLR were noted between AECOPD and stable group. It might suggest that the sub-phenotype of neutrophils, rather than its total number, was responsible for the alteration of NLR. Our results were also consistent with those findings by previous study that NLR was an independent risk factor for patients with COPD with exacerbations, but the potential mechanism remain poor understand(44). This study proposed the critical involvement of PD-L1-PD-1 axis in mediating AECOPD by mediating NLR. These findings were further confirmed by significant correlation between the expression of PD-L1 on neutrophil and FEV1%, which was also conformed with previously reported correlation between NLR and FEV1%(5).

In our study, the cutoff value for predicting AECOPD is 4.31, which was similar with other studies that found NLR values over 3.29(45) and 3.34(46) were able to predict AECOPD. PD-L1 expression was the only statistically significant predictor of AECOPD in regression analysis, exhibiting the highest area under ROC curve in current study. We found that a PD-L1 cutoff value of 3.273 serves as a predictive marker for the occurrence of AECOPD. This finding holds significant implications for clinicians; specifically, when the levels of PD-L1-expressing neutrophils in patients with COPD exceed 3.273, clinicians should be vigilant regarding the potential risk of acute exacerbation. Consequently, it is advisable to proactively adjust the patient’s medication based on this predictive value to mitigate the risk of adverse outcomes. Given the positive correlation between PD-L1 expression and NLR, it is reasonable that PD-L1 expression on neutrophil is of great significance in predicting AECOPD by regulating NLR, hinting that the expression of PD-L1 on neutrophil was more powerful marker to predict AECOPD.

## Limitation

This study had certain limitations. Firstly, this study is conducted at a single center and involves a relatively small sample sizes, which needs for future multi-center replication studies. Secondly, some other outcomes, such as long-term control of symptoms, activity of daily life, and mortality also remain unclarified due to small sample sizes and short-term follow-up. Thirdly, this study did not confirm direct contact between neutrophil PD-L1 and lymphocytes, although increased neutrophil PD-L1 expression was associated with lymphocytes; however, the mechanisms need further validation. Further functional studies are required to confirm the role of PD-L1 role. Lastly, our study focused on the Han Chinese population. Future research should consider incorporating participants from diverse countries and ethnic backgrounds to enhance the study’s generalizability.

## Conclusion

Our studies provide evidence that neutrophil PD-L1 serves as an independent risk factor in predicting the incidence of AECOPD, which might be due to disrupting NLR and worsening FEV_1_%. Experimental and longitudinal studies are required to further validate these findings.

## Data sharing statement

The data that support the findings of this study are available from the corresponding author upon reasonable request.

## Data Availability

All data produced in the present study are available upon reasonable request to the authors

## Acknowledgment

### Financial/nonfinancial disclosures

National Natural Science Foundation of China 82272187, 81801935 ; Beijing Hospitals Authority Youth Programme (No. QML20230309); Reform and Development Program of Beijing Institute of Respiratory Medicine; Beijing Physician Scientist Training Project BJPSTP-2024-24 Financial Budgeting Project of Beijing Institute of Respiratory Medicine(No. Ysbz2025004).

## Author contributions

Chao Ren had full access to all the data in the study and take full responsibility for the integrity of the data and the accuracy of the data analysis. Wei Sun contributed substantially to the study design, data and interpretation, and writing of the manuscript. Jing Wang contributed substantially to the study design, data and interpretation. Liyu Zheng contributed to blood neutrophil isolation and flow-cytometry analysis. Haibin Li was responsible for the statistical analysis. Li An contributed to the background investigation and study design.

